# Excess cases of Influenza like illnesses in France synchronous with COVID19 invasion

**DOI:** 10.1101/2020.03.14.20035741

**Authors:** Pierre-Yves Boëlle, the Sentinelles syndromic and viral surveillance group

## Abstract

Several French regions where COVID19 has been reported currently show a renewed increase in ILI cases in the general practice based *Sentinelles* network. Here we computed the number of excess cases by region and found correlation with the number of reported COVID19 cases so far. These data suggest larger circulation of SARS-CoV-2 in the French population than apparent from confirmed cases.

## Background

The *Sentinelles* network monitors Influenza Like Illnesses (ILI) and Acute Respiratory Infections (ARI) in general practice in France(1). According to routine surveillance, the 2019-2020 influenza epidemic reached its peak in mid-February in France (http://www.sentiweb.fr) and it was expected that ILI incidence would decrease over time, as it did over all past seasons.

COVID19 was first identified in France in the end of January(2). The number of confirmed cases reported to the health authorities was 979 by March 7, 2020. At the same date, we found out that the number of ILI cases reported to the Sentinelles network showed a renewed increase in some regions. Furthermore, several swabs collected by Sentinelles GPs tested positive for SARS-CoV-2.

Here, we aimed at quantifying the number of consultations for ILI in France in early March 2020 in excess of what was expected due to the influenza epidemic and to examine its relationship with reported cases of COVID19.

## Methods

Influenza Like illnesses cases are defined as cases with fever of sudden onset (>39°C) with respiratory signs (cough, running nose) and myalgia of any age. Acute Respiratory Infection (AIR) include diseases with respiratory signs and are only monitored in the > 65 y.o. Cases are reported in real time by participating GPs (approx. 600 GPs) all over France. Additionally, *Sentinelles* GPs may swab 1 ILI case and up to 2 IRA cases per week for viral characterization(3). Influenza virus, respiratory syncytial virus, human rhinovirus and human metapneumovirus are routinely looked for, and SARS-CoV-2 has been added since February 2020.

We computed the expected number of ILI consultations using the superposition of a seasonal (4) and an influenza epidemic component(5), as detailed in the appendix. Excess cases were computed as the difference between observed cases and expected numbers. The rate of increase in excess cases was determined, assuming exponential growth.

Confirmed COVID19 cases were obtained from the santé Publique France website, March 7 Bulletin(6).

## Results

### SARS-CoV2 positive swabs in ILI and ARI cases

In week 10 of 2020, 93 swabs were collected in ILI patients and 23 in ARI patients. Two (2) swabs in each category were positive for SARS-CoV-2. The 2 positive swabs in ARI patients had been collected in In Bourgogne-Franche Comté (BFC) and Grand Est (GRE) regions, out of a total of 7 swabs in these 2 regions. Assuming a positivity rate for SARS-CoV-2 of 2/7 in these regions, we estimated that 760 (CI95% [219, 1706]) ARI consultations in the > 65 y.o. could have been caused by COVID19 during week 10 (2/7 of 2600 ARI visits). One of the positive ILI case was linked to an existing cluster in the east of France.

### Analysis of ILI incidence

The modelling approach allowed computing excess cases in 11 out of 13 regions. The model did not provide a good fit in the remaining 2 regions (see appendix). The overall ILI incidence showed renewed increase with 33 [-8, 64] consultations/100000 in excess during week 9 and 84 [447, 108] consultations/100000 in excess in week 10. This changed with regions: 4/11 regions displayed positive excess (credible interval excluding 0) in week 9 and 7/11 regions in week 10.

**Figure 1:**
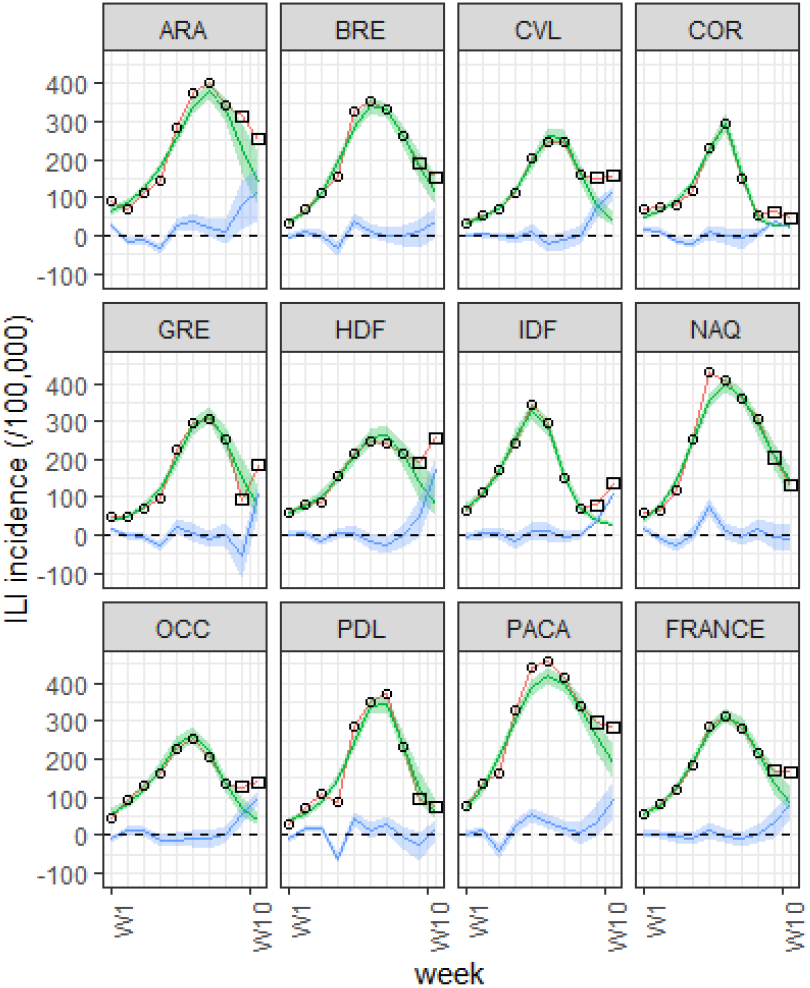
Number of consultations for ILI (per 100000) in France and 11 French regions from 2020 W1 to 2020 W10, with expected number of consultations (gren) fitted on the first 8 weeks of 2020 (circles). Excess consultations (blue) computed by difference between observations (squares) and expected counts. See Table1 for region codes.

The number of excess cases correlated with the cumulated number of COVID19 cases reported in the same regions (r=0.61, p<0.05).

The weekly exponential increase rate estimated from week 2020/8 to 2020/10 was 0.69 (CI95% [0.55, 0.86]) in GRE, 0.67 (CI95% [0.55, 0.83]) in IDF, 0.68 (CI95% [0.56, 0.83]) in HDF, 0.61 (CI95% [0.48, 0.75]) in OCC and 0.56 (CI95% [-1.6, 0.8]) in France overall.

**Table 1:** Excess consultations/10000 in French regions in 2020 week 9 and 10 and cumulated number of COVID19 cases by March 7.

| Region | Consultations in excess(/100000) |  | COVID19 Cases (March 7) |
| --- | --- | --- | --- |
|  | Week 9 | Week 10 |  |
| Auvergne-Rhone-Alpes (ARA) | 82[17, 147] | 115 [36, 174] | 102 |
| Bretagne (BRE) | 9 [-32, 44] | 40 [-7, 73] | 40 |
| Centre-Val de Loire (CVL) | 72 [43, 96] | 117 [97, 130] | 16 |
| Corse (COR) | 35 [32, 37] | 21 [20, 23] | 5 |
| Grand-Est (GRE) | -55 [-109, -13] | 107 [56, 138] | 250 |
| Hauts de France (HDF) | 51 [-3, 94] | 172 [111, 208] | 173 |
| Ile de France (IDF) | 40 [32, 46] | 107 [103, 110] | 104 |
| Nouvelle Aquitaine (NAQ) | -3 [-110, 32] | -9 [-44, 26] | 17 |
| Occitanie (OCC) | 54 [30, 75] | 99 [79, 112] | 36 |
| Pays de la Loire (PDL) | -25 [-70, 5] | 18 [-16, 38] | 18 |
| Provence-Alpes-Cote d'Azur (PACA) | 36 [-4, 80] | 94 [47, 140] | 38 |
| France | 33 [-8, 64] | 84 [44, 108] | 939 |

### Interpretation

In the last 30 seasons of routine surveillance in France with the Sentinelles network, this is the first time that an increase in ILI cases is observed simultaneously in several regions after the peak of the annual influenza epidemic.

Several processes may be at play in this observation, including at least characteristics of the influenza season, change in population behaviour or increase in COVID19 incidence.

H1N1pdm and B influenza viruses co-circulated in 2019-20 in France. Seasons with the type B virus can lead to prolonged periods for decrease in incidence but a renewed increase has never occurred before (1). The school holidays occurred gradually in France during the end of February and early March. While this may change the dynamics of influenza(7), the effect is generally notable in the ascending phase of the epidemic rather than after the peak.

The current situation with COVID19 may change the health-seeking behaviour of patients and, to some extent, of the contributing general practitioners although they conform to a case definition. Preliminary assessment of behavioural data in crowdsourced surveillance system is under way (grippenet.fr(8)) but, as of today, no massive trend towards more consultations has been observed. As information regarding the coronavirus risk is widespread, a uniform increase over all regions could have been expected in this scenario, but we acknowledge that it may have led to increased consultation rates in regions where COVID19 is the most reported.

The overlap between COVID19 and flu symptoms makes it possible that the excess of ILI cases is due to COVID19 cases, and the presence of SARS-CoV-2 positive swabs in the patients demonstrate this possibility. The correlation between the number of confirmed cases and the computed excess is also consistent with this scenario. In this case, the excess in cases is compatible with an exponential growth with rate estimated at approximately 0.7/week. Interestingly, this estimate is very similar from one region to the next in the four most affected regions (GRE, IDF, OCC, HDF). This estimate may prove useful to help calibrate models for studying the impact of the COVID19 pandemic.

Estimating the true number of COVID19 cases remains difficult in the presence of these uncertainties. We note that the extrapolated number of COVID19 presenting as ARI was 760 in the >65 y.o. in the 2 eastern regions (GRE and BFC), which was at least twice more than the number of reported cases at the time (250+129), all ages confounded.

As we enter a period of generalized circulation of the SARS-CoV-2 virus, surveillance based on clinical description and swabbing by GPs will prove essential to help us assess the situation.

## Data Availability

data available from http://sentiweb.fr and other public websites.

To ensure timely distribution of the note, authors are presented as “the Sentinelles syndromic and viral and surveillance group” including (but not limited to) Cécile Souty^1^, Titouan Launay^1^, Clément Turbelin^1^, Caroline Guerrisi^1^, Chiara Poletto^2^, Vittoria Colizza^2^, Sylvie van der Werf^4,5,6,7^, Bruno Lina^8,9^, Daniel Lévy-Bruhl^3^, Thomas Hanslik,^1^ Thierry Blanchon^1^

1 – Sorbonne Université, Institut Pierre Louis d’Epidemiologie et de Santé Publique, Paris, France

2 – INSERM, Institut Pierre Louis d’Epidemiologie et de Santé Publique, Paris, France

3 – Santé Publique France, Saint Maurice, France.

4- Institut Pasteur, Unité de Génétique Moléculaire des Virus à ARN, Paris, France

5- Institut Pasteur, Centre Coordonnateur du Centre National de Référence des virus des infections respiratoires (dont la grippe), Paris, France

6- UMR CNRS 3569, Paris, France

7- Université Paris Diderot, Sorbonne Paris Cité, Unité de Génétique Moléculaire des Virus à ARN, Paris, France

8- Laboratoire de Virologie, Hospices Civils de Lyon, Institut des Agents Infectieux (IAI), Centre National de Référence des virus respiratoires (dont la grippe), Centre de Biologie et de Pathologie Nord, Groupement Hospitalier Nord, Lyon, France

9- Université de Lyon, Virpath, CIRI, INSERM U1111, CNRS UMR5308, ENS Lyon, Université Claude Bernard Lyon 1, Lyon, France

## Appendix

The number of expected ILI consultations reported to the *Sentinelles* network was modelled as the superposition of 2 components: an “epidemic” component due to influenza infections and a background seasonal signal not due to influenza (4). More precisely, we fitted the influenza epidemic part using Richard’s model(5). Richard’s model is a 4 parameters equation describing the cumulated number of cases at time *t* as follows :

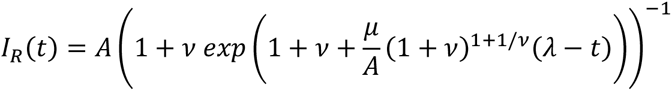

The seasonal part was modelled as

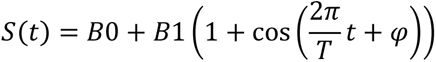

We fitted this model assuming that the number of ILI consultations ILI(t) was Poisson distributed with mean *I*_*R*_(*t*)+*S*(*t*)-over the 8 first weeks of 2020 and used non-epidemic periods in years 2016 to 2020 additional information to estimate *S*(*t*)-(see appendix for more detail). Excess ILI cases was defined as observation minus the expected part, i.e. *E*(*t*)=*ILI*(*t*) - *I*_*R*_(*t*) - *S*(*t*).

In a second step, we fitted excess cases by an exponential increase *C*_*E*_(*t*)=*C*0 (-*t*), and assumed that number of ILI cases for weeks 8 to 10 was Poisson distributed with mean *I*_*R*_(*t*)+*S*(*t*)+*C*_*E*_(*t*). We tied the number of excess cases at 0 in week 1 of 2020 for this estimation.

Estimations were performed with STAN. Code is available upon request.

Example data used for estimating excess cases in a French region. The orange points were used to estimate the seasonal part; the first peak in the black part was fit using Richard’s model plus the seasonal part, and excess was computed over the last 2 weeks as indicated in figure S3.

**Figure S1:**
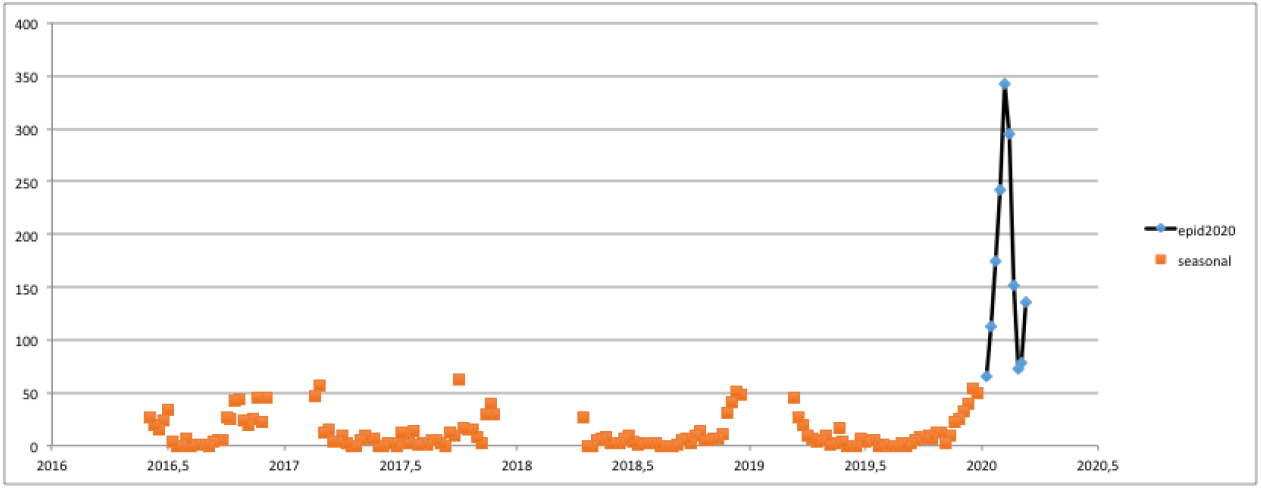
Example data used for excess consultations modelling. The black line corresponds with the 2020 epidemic. The orange points corresponds with the seasonal background information from previous years.

**Figure S2:**
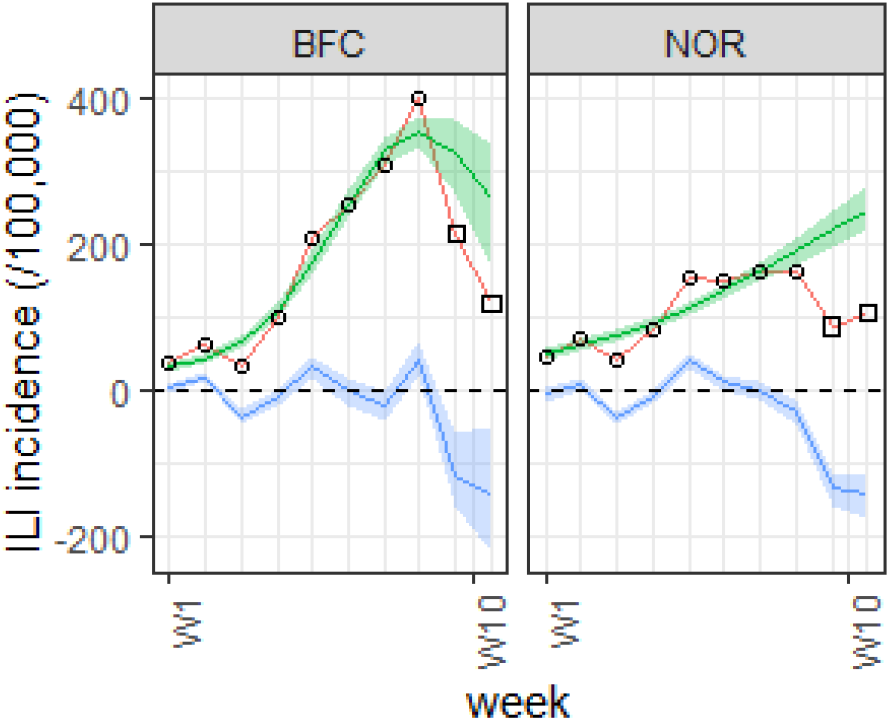
Two French regions where the model failed to describe the data.

**Figure S3:**
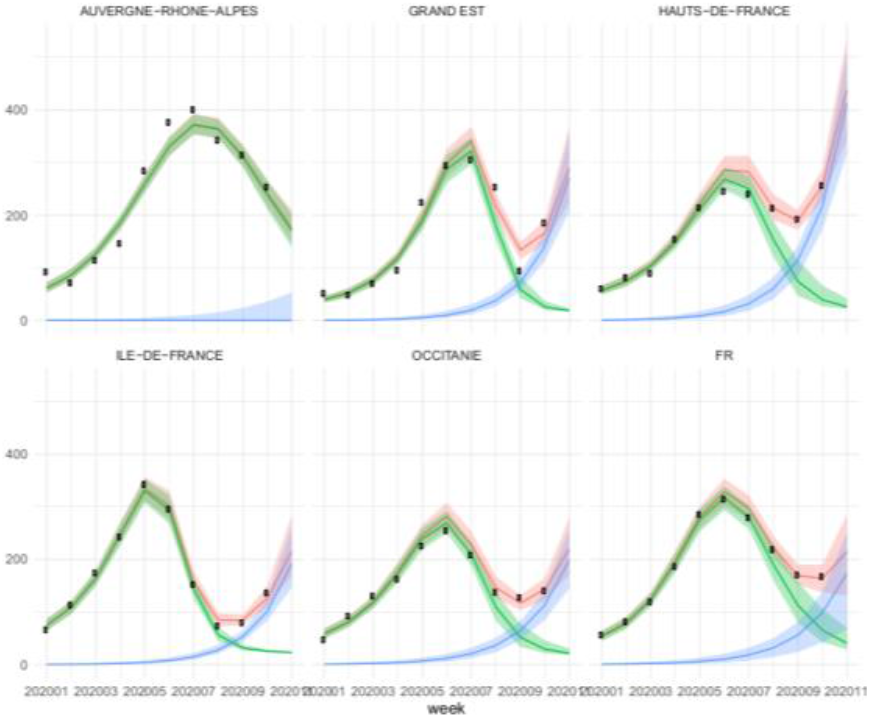
Decomposition of observed ILI cases in France (dots) in a seasonal + epidemic part (green) and excess part (blue). The red curve is the sum of the two components.

